# Measuring progress on health and well-being in the Eastern Mediterranean Region via voluntary national reviews, 2016 - 2021: what does the data reveal?

**DOI:** 10.1101/2024.01.02.24300730

**Authors:** RM Mabry, HV Doctor, MN Khair, M Abdelgalil, A Rashidian

**Author notes:** **Corresponding Author:** Ruth M. Mabry, Global Health Consultant, Muscat, Oman.

## Abstract

**Background:** Country **s**ubmission of Voluntary National Reviews (VNRs) is the formal mechanism to report on progress of the Sustainable Development Goals (SDGs). Despite strong political commitment to strong information systems, large data gaps exist in the Eastern Mediterranean Region.

**Methods:** This study aims to review VNRs submitted by countries in the region to assess the comprehensiveness of reporting on the health-reported SDG targets and indicators. We conducted a content analysis of VNRs of 18 countries of the region submitted between 2016 and 2021. The review focused on progress on the SDGs by assessing i) the reporting on the 50 health-related targets and indicators ii) data availability using the WHO reporting framework, and iii) data availability based on source of information. Spreadsheets were developed and used to extract data and facilitate content analysis.

**Results:** All VNRs confirmed that SDG monitoring and reporting mechanisms have been established, however, only 11 VNRs reported on all 17 SDGs and 8 explicitly mentioned country specific 2030 targets. Many VNRs identified data availability as a key challenge to SDG monitoring; for the health SDG, data availability ranged from 48% to 93% among the five countries reporting this figure. Comprehensiveness of reporting varied by type of indicator (maternal, child and infant mortality were the most common) and by country income level (greater reporting by high income countries).

**Conclusions:** Significant work remains to enhance information systems across the region to monitor progress and guide actions to achieve the health-related SDGs. Strengthening health information systems regulatory frameworks, data collection capacities including strengthening civil registration and vital statistics and population-based surveys are key steps to enhancing access to quality data which in turn can contribute to achieving the health-related SDGs.

## Introduction

In 2015, the United Nations (UN) General Assembly adopted the 2030 Agenda on Sustainable Development (1). Building on the Millennium Development Goals, this new agenda outlined an ambitious plan addressing 17 Sustainable Development Goals (SDGs) and 169 targets to be achieved by 2030. Countries regularly prepare Voluntary National Reviews (VNRs) to the United Nations High-level Political Forum on Sustainable Development, the formal mechanism to report on progress and share lessons learned on SDG implementation (2).

SDG3 on health is a key indicator of success in achieving the 2030 Agenda (3). Progress on the health-related SDGs is slow around the world.(4) Prior to the COVID-19 pandemic, progress was made on only half of the 50 indicators in the Eastern Mediterranean Region (EMR); the rate of progress on seven of them is too slow to meet global targets (5, 6). By 2023, progress is nearly at a standstill with marked setbacks across most indicators on health, health risks and determinants and access to health services in the region (7).

Access to quality data can contribute to achieving the health-related SDGs; better data and information is essential for evidence-informed decision-making, proper management and rational resource allocation and for monitoring and reporting (8, 9). Quality reliable data requires a well-functioning national health information system. However, limited availability of data in countries of the regions, especially lack of trends and disaggregated data – one in five indicators are not available and one in four indicators are from 2019 or earlier – is a major challenges for monitoring progress (7). Despite, strong political commitment to having national health information systems that respond to national needs, large data gaps constrain the ability for policy-makers to take evidence-based public health actions (6, 10–12).

A regional reviews of 60 VNRs submitted by countries in the European Region reported weak national and subnational health information systems as a core challenge (13). Similarly, a review of VNRs submitted by 18 countries in the Eastern Mediterranean Region identified data availability as one of the most common challenges to SDG implementation (14). Building on this regional review, this study aims to assess the comprehensiveness of reporting on the health-reported SDG targets and indicators and reflect on country-level priorities for monitoring them.

## Methods

Similar to our earlier review (14), here we reviewed the latest VNRs from 18 countries of the EMR submitted between 2016 and 2021 and available on the UN website.^a^ The review focused on progress on the SDGs, especially the health-related ones. Tracking progress involved assessing the reporting on the 50 health-related SDG targets and indicators; including 32 indicators associated with the 17 SDG3 targets. We also assessed data availability using the WHO reporting framework of 47 indicators across 5 domains (mortality, morbidity, means of implementation, risk factors and determinants) (6), comparing the availability of data as reported by WHO Regional Office for the Eastern Mediterranean (EMRO) for the same year as the VNR. These data are provided and validated by countries annually. We also assessed availability of data based on source of information recognizing that well-functioning health information systems would be able to provide the necessary information to report on 29 of the 32 SDG3 indicators: civil registration and vital statistics system (11), country national survey (6), the disease surveillance system (3) and the routine health information system (9) (Supplementary Table 1) (8).

Spreadsheets were developed and used to extract data and facilitate content analysis in terms prioritization of SDGs and reporting of SDG health-related indicators. The data that support the findings of this study are available on request from the corresponding author.

## Results

All 18 VNRs reviewed indicated that mechanisms have been established to monitor and report on progress on the SDGs. Eleven VNRs reported on all 17 SDGs, the remaining reported on seven to 12 SDGs (Figure 1). The comprehensiveness of reporting across SDGs was associated with country income levels so that higher income countries tended to report on more SDGs. 17 VNRs reported that targets and indicators had been prioritized and adapted to the country context. However, only 8 VNRs explicitly mentioned country targets for 2030 for at least some of the indicators. Egypt, for example, identified SDGs “Push” targets across all the SDGs. Morocco described the level of advancement for each target and explained their improvement plan. Libya mentioned the targets for the 10 SDGs for which they reported on. Other countries had a more limited approach. For example, Jordan identified targets for SDGs 1, 6, 7 and 13; Lebanon only mentioned targets for SDG 6, 7, 13 and 15. Pakistan grouped the SDGs into 3 categories; SDGs for immediate attention (2, 3, 4, 7, 8, and 16) identified specific targets.

**Figure 1.**
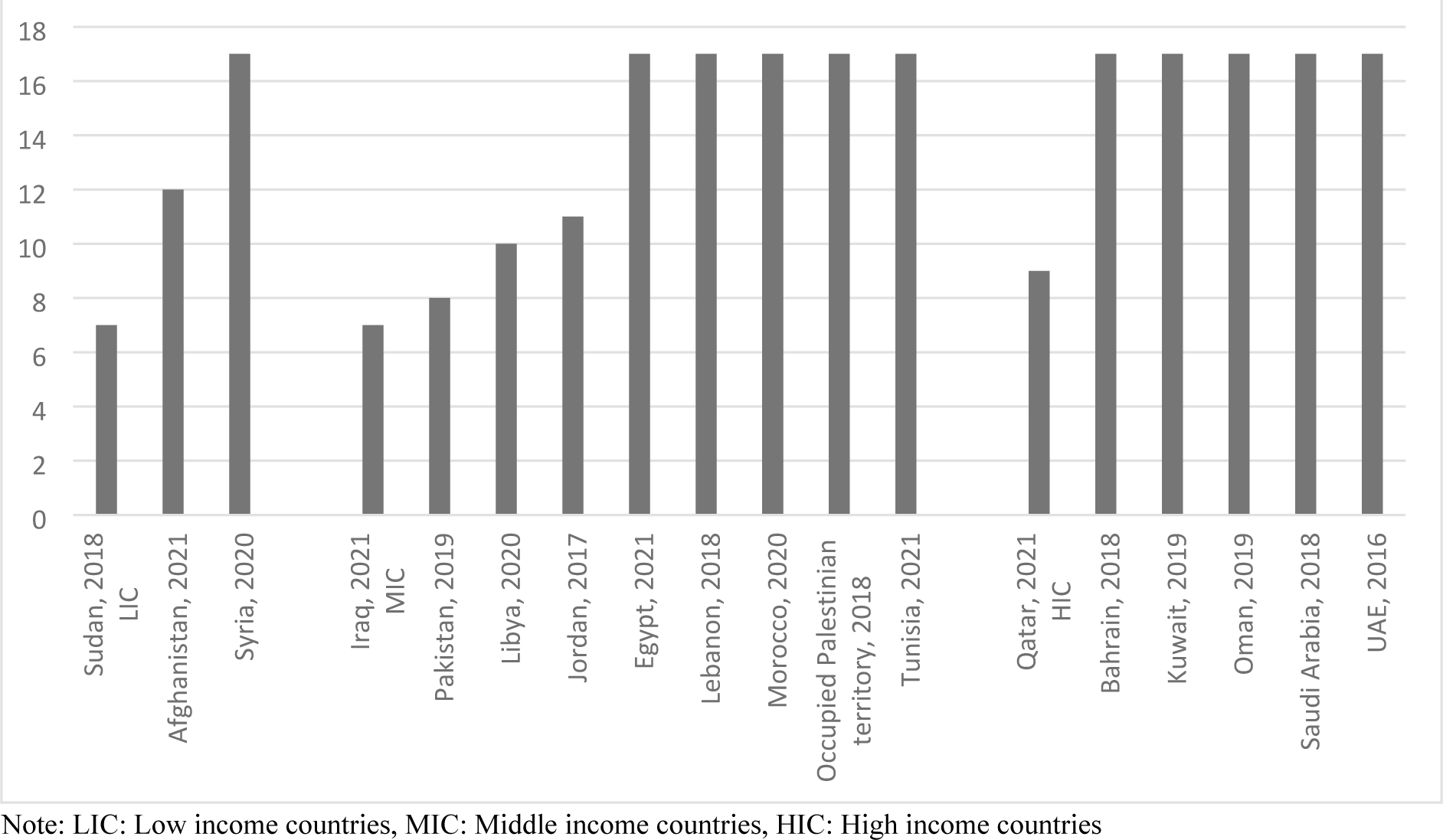
Number of SDGs prioritized by countries in their VNRs (N=18)

Nine VNRs (Afghanistan, Egypt, Iraq, Jordan, Morocco, occupied Palestinian territory, Oman, Pakistan, and Qatar) reported on the data availability (Table 1). Of the seven countries (Egypt, Iraq, Jordan, Morocco, occupied Palestinian territory, Oman, and Qatar) that reported results of data gap analyses, Morocco and Qatar, were the only ones reported more than 50% data availability for the 244 SDG indicators. The availability of data for SDG3 ranged from 48% to 93% among the five countries (Jordan, Morocco, Oman, occupied Palestinian territory, and Qatar) reporting this figure with Oman was the only country which reported higher availability of data for SDG3 than for all the other Goals. Fourteen VNRs mentioned limited data availability as one of the challenges to monitoring and reporting. They usually described the limitations in terms of national human resource capacity to collect and/or analyse information. Additional challenges mentioned included low data quality, lack of statistical tools, limited transparency in data dissemination, and lack of data disaggregation.

**Table 1.**
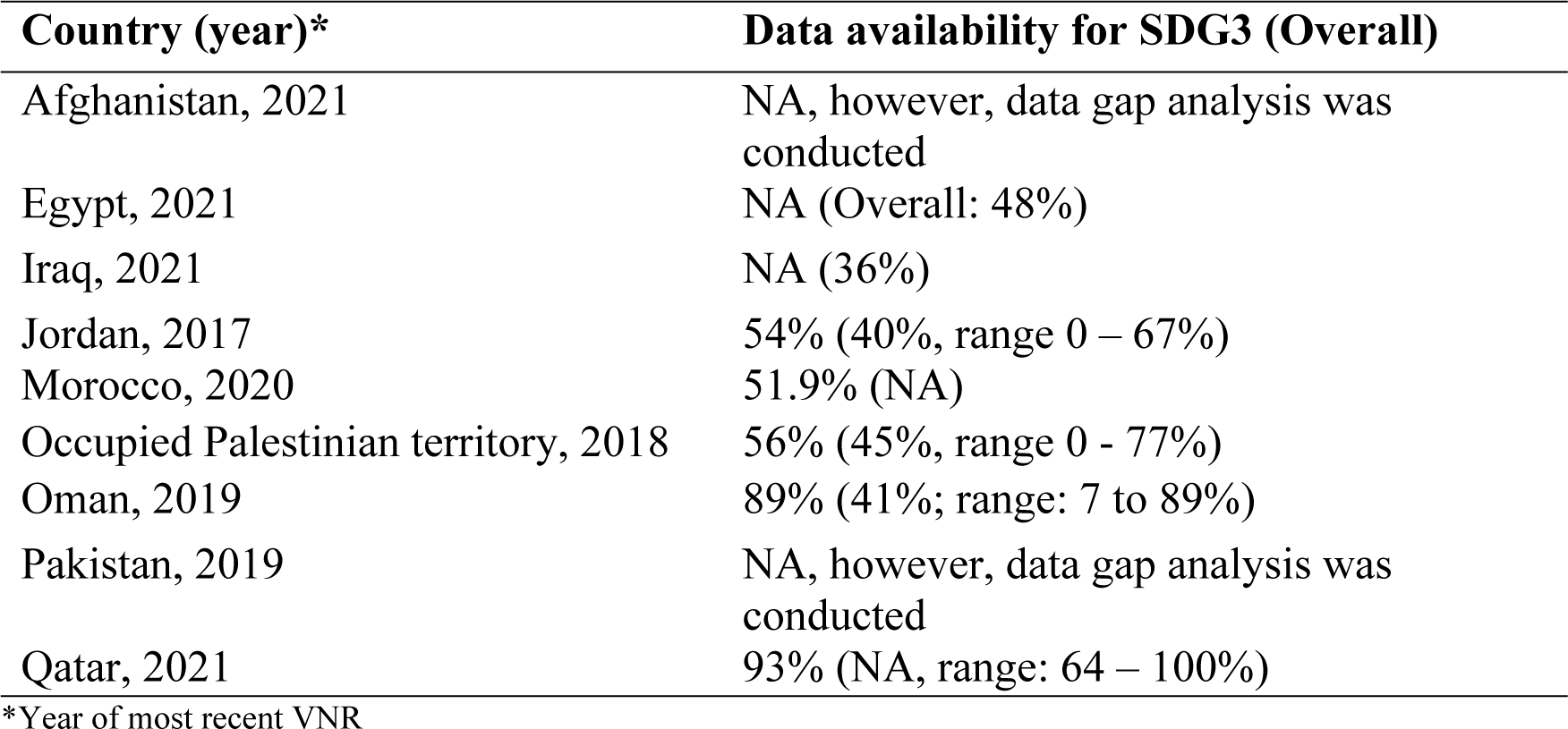
Data availability as described in VNRs, EMR.

| **Country (year)\*** | **Data availability for SDG3 (Overall)** |
| --- | --- |
| Afghanistan, 2021 | NA, however, data gap analysis was conducted |
| Egypt, 2021 | NA (Overall: 48%) |
| Iraq, 2021 | NA (36%) |
| Jordan, 2017 | 54% (40%, range 0 – 67%) |
| Morocco, 2020 | 51.9% (NA) |
| Occupied Palestinian territory, 2018 | 56% (45%, range 0 - 77%) |
| Oman, 2019 | 89% (41%; range: 7 to 89%) |
| Pakistan, 2019 | NA, however, data gap analysis was conducted |
| Qatar, 2021 | 93% (NA, range: 64 – 100%) |

\*Year of most recent VNR

The comprehensiveness of reporting progress on specific health-related indicators varied by type of indicator and by country income levels. The highest proportion of indicators reported were by high income countries (HICs) for mortality and morbidity indicators (60%); the lowest were the means for implementation reported by low-income countries (LICs) and middle-income countries (MICs); 14.7% and 20.6%, respectively. MICs and HICs reported a higher percentage of targets across the five areas compared to LICs. Unlike with disease indicators, MICs reported on more targets in the areas of risk factors (50.0% and 35.7%, respectively) and determinants compared to HICs (52.1% and 35.4%, respectively) (Figure 2).

**Figure 2.**
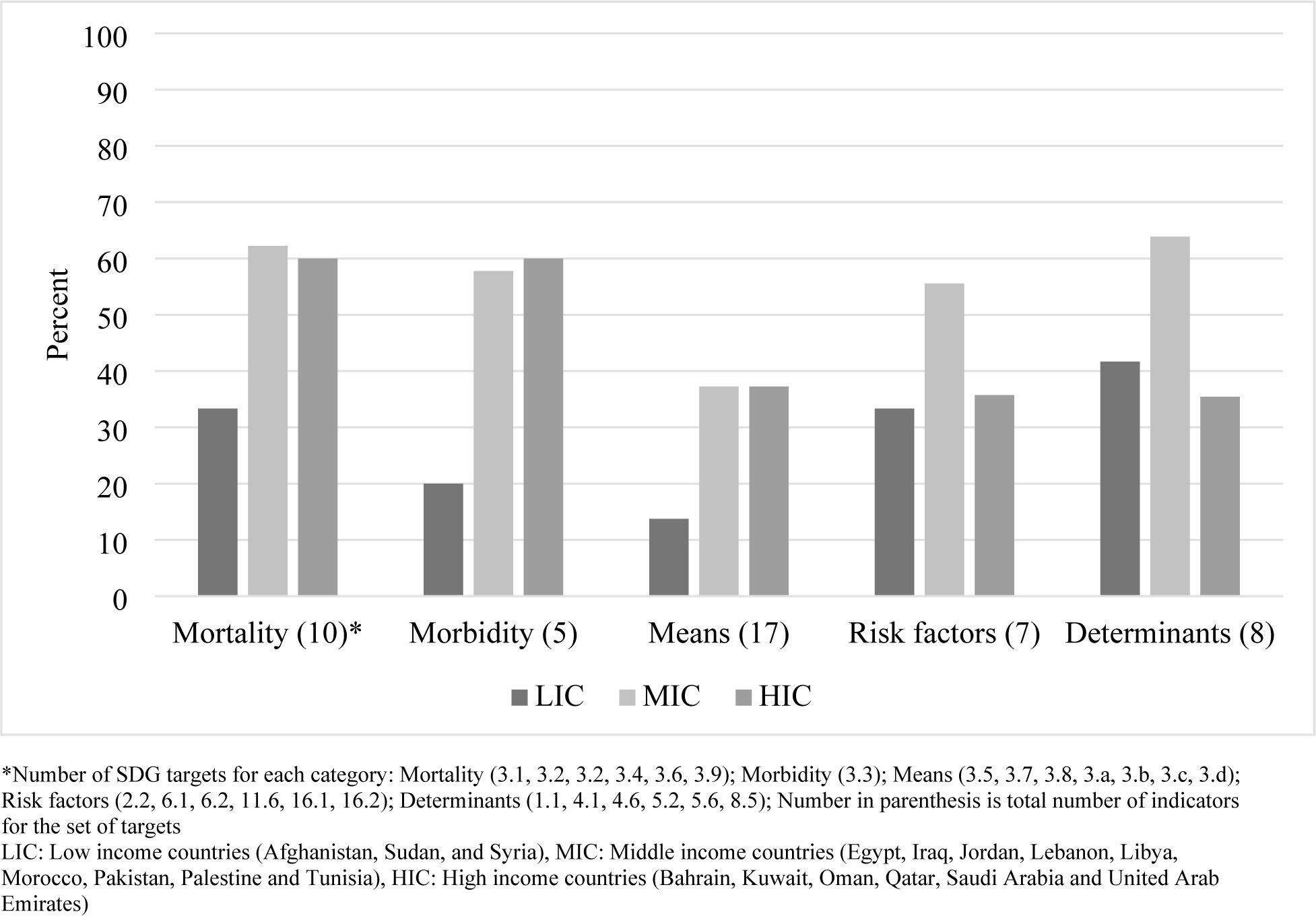
Percent of reported SDG health-related indicators in EMR Voluntary National Reports, by income levels.

The most commonly reported indicators were those related to SDG3 including 3.1.1. maternal mortality ratio (100%), 3.2.2 neonatal (83%) and 3.2.1 under-5 mortality rates (78%) (Figures 3 and Supplementary Tables 1 and 2). The other indicators that reported by at least 70% of the VNRs included access to 6.1.1 improved drinking water (78%), 3.3.1 new HIV infections (72%) and 8.5.2 total unemployment rates (72%). Reporting on progress with at least two data points was less common; the most common indicator for reporting trends figures was for maternal mortality ratio (50%).

**Figure 3.**
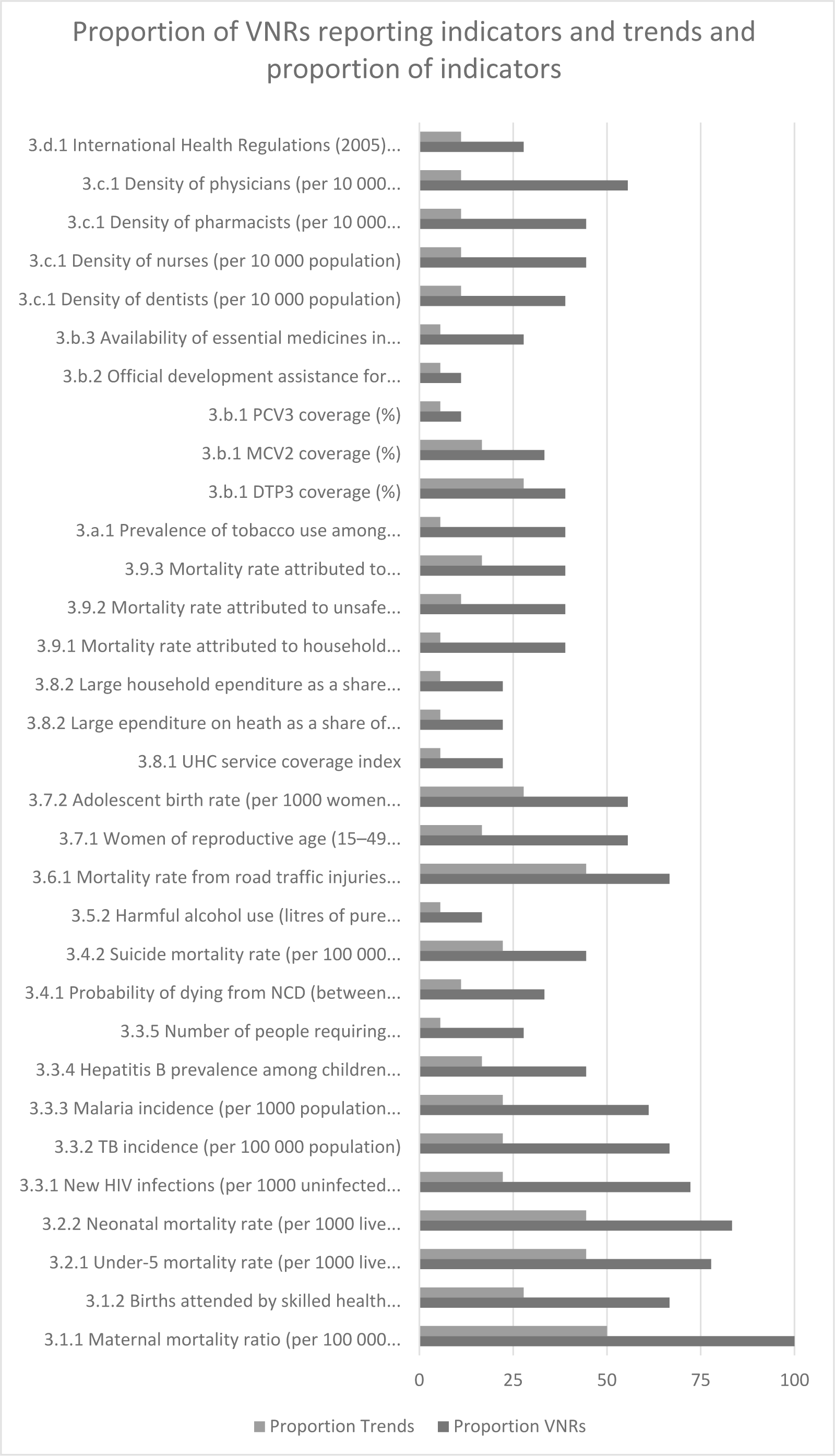
Proportion of VNRs reporting SDG3 indicators and trends.

Qatar was the only country that reported on trends across all the SDG3 indicators; all other countries provided trend data for 9 or less SDG3 indicators (Supplementary Table 1). On average, the VNRs provided data for about 11 indicators linked to SDG3 targets (Supplementary Table 1) and 6.4 indicators to non-SDG3 indicators (range 0 – 12, Table 2). Data available from the WHO/EMRO reports (Supplementary Tables 1 and 2) often were not included in the VNRs even though these figures are routinely reported to WHO/EMRO by countries.

Twenty-nine of the 32 SDG3 indicators can be derived from four key data sources of a well-functioning health information systems: civil registration and vital statistics (CRVS) system (11 indicators), country national survey (6), the disease surveillance system (3) and the routine health information system (9) (Supplementary Table 1). The remaining three, harmful alcohol use, official development assistance and IHR capacity are obtained from country administrative data, the Organization for Economic Cooperation and Development and key informant surveys and not necessarily from within the national health information system. The reporting of the health-related indicators in the VNRs was much higher for the indicators derived from the disease surveillance system (66.7%) and CRVS system (67.8%) compared to those based on routine health information system (41.1%) and national surveys (36.7%, Figure 4).

**Figure 4.**
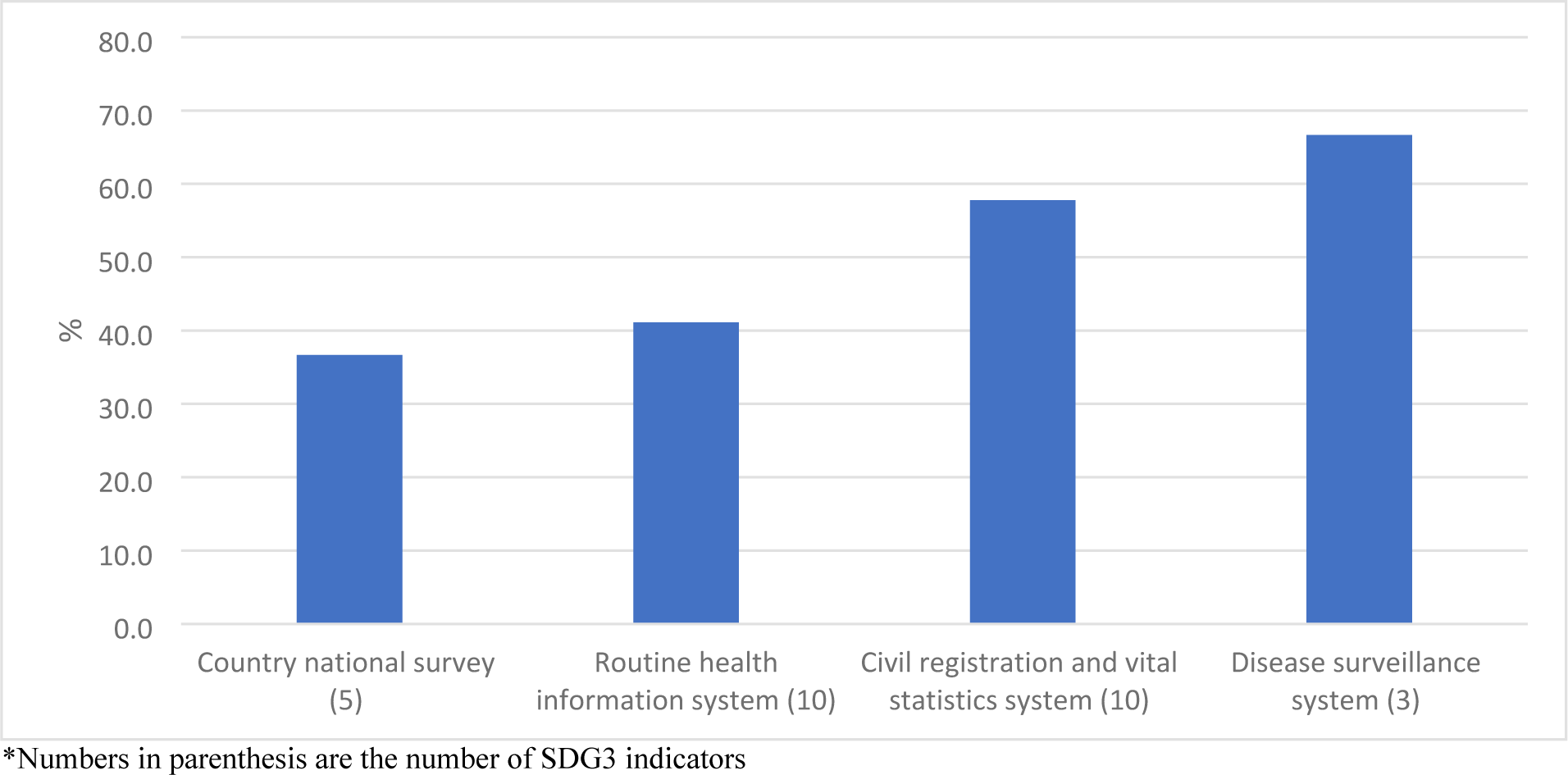
Percentage of SDG3 indicators reported in VNRs by data source.

## Discussion

This review of VNRs from 18 countries of the EMR confirmed countries’ commitment to monitor progress on the SDGs with two of three reporting across all goals, particularly countries of higher income levels. The comprehensiveness of reporting progress on specific health-related indicators varied by type of indicator and by country income levels with maternal, neonatal and under-5 mortality rates the most commonly reported indicators. Limitations to monitoring progress on indicators were described in terms of national human resource capacity to collect and/or analyse information with a few mentioning issues like low data quality, lack of statistical tools, limited transparency in data dissemination, and lack of data disaggregation. Findings demonstrate capacity gaps: high proportion of data unavailable to measure progress, limited presented of data for reporting progress and only one country reporting trends across all SDG3 indicators. Given the importance of access to quality data to achieving the SDGs, significant work remains to ensure well-functioning national health information systems in countries in the region (8, 9).

All VNRs demonstrated commitment to a multisectoral approach to collecting data by appointing a national authority for this task. Having a central authority is a key component of a strong governance structure but it is not enough. Achieving universal health coverage requires universal access to essential health information (15, 16). A regional review of national health information systems found an inadequate regulatory framework across the 10 countries assessed (17). Legislation, regulations, policies, strategies and standards that establish clear roles and responsibilities to all involved promote data integration of different information system, and regulate access to the information (18). It also requires increasing investment in health information systems. Adopting an open access approach to sharing data can make health related data collected by various sectors more widely available and maximize their usage for policy-making (9, 19). Moving forward developing an investment framework defining the roles and responsibilities of relevant sectors to avoid duplication and updating legislations and policies is a key step to improving functionality of national health information systems (17, 18, 20).

VNRs are highly formalized comprehensive reports; including an annex with data with gender statistics and disaggregated data, is encouraged but not required (2). Being voluntary, these progress reports providing details of specific targets and indicators for all 17 SDGs is not mandatory which would be a key reason why limited data is included in the reports. Additionally, as strategic documents, varying details on efforts made in data availability, quality, and coverage is understandable. Reporting at a high strategic level without providing comprehensive reporting on over 150 indicators does not necessarily indicate inability to report on specific indicators. On the other hand, data availability is a known challenge for monitoring regional progress on the health-related SDGs; for although two or more data points are available for a good portion of the indicators reported to WHO, the latest data for around 20% is four years or older (7). This challenge is not unique but was also reflected in the VNR review conducted in Europe along with concerns about limited analytical capacity, poor investment in technology and systems, and weak monitoring and evaluation (13). Similarly was the lack of disaggregated data across all VNRs in this review like the review of VNRs from Europe (13).

Comprehensiveness of reporting was similar between middle-and high-income countries for mortality, morbidity, and means related indicators with the highest proportion being only 60%; middle income countries also had higher levels of reporting for risk factors and determinants indicators. Regionally, evidence indicates that barely two-thirds of births are registered, slight over half of death are registered, where six of 22 countries have sustainable capacity for public health surveillance and barely half have conducted at least one survey within the last 10 years (the lowest among all WHO regions) (21). Collecting a small set of locally-relevant and policy-oriented indicators is key for successful implementation of health plans; this requires engaging across sectors to improve statistical capacity and increase data quality and availability (6, 12, 22). Countries should focus their efforts in building national capacity across all data sources, particularly CRVS and population-based surveys.

CRVS systems are a foundation for good governance; birth and death registration are essential for estimating disease burden and for assessing the impact and cost-effectiveness of public health programmes and health interventions (23). Ten of the 50 regional core indicators depend on a well-developed CRVS system. Although some progress has been made in completeness of vital event registration since 2015 when the region endorsed a strategy to strengthen their CRVS system (24, 25), a large portion of births and deaths remain unregistered (26). Various policy approaches that include both supply and demand components based on country’s health system governance and sociocultural context, can improve registration. Increasing public awareness on the importance of birth and death registration, introducing electronic death notification platforms and digital notification systems, and periodic evaluations of the quality of medical certification of causes of death, are some of the priority actions recommended by WHO to improve this system in countries of the region (18). Documenting national policies that help strengthen systems in countries of the region would provide useful evidence in shaping regionally relevant policies and operational considerations for strengthen CRVS systems in the EMR (23).

National population-based surveys are critical components for a health information system supportive for SDGs; particularly so countries can have locally derived information rather than depend on international estimations (12). They complement CRVS systems as a key source of information to monitor trends and assess inequality. A large majority of the regional core indicators could be generated from national population-based surveys including the health examination survey, Demographic Health Survey (DHS), Multiple Indicator Cluster Surveys (MICS) and the Global School Health Survey (27). These surveys, conducted over a 10 year period, could generate more robust data than available from routine systems currently available in the region (27). Developing and implementing a national survey plan would enable a country to obtain a good portion of health-related SDG3 indicators (18).

VNRs provide a comprehensive overview of progress on the SDGs where reporting on targets on indicators is voluntary; their limitation inclusion does not necessarily indicators a country’s inability to report on them. At the same time, the VNR preparation process results in a lengthy report with few if any references so interpretation is left of a subjective judgement of the authors.

## Conclusion

VNRs from the countries in the EMR while demonstrating commitment to the SDGs recognized weak national health information systems, limited data availability and analytical capacity as challenges to monitoring and reporting. Strengthening health information systems regulatory frameworks, data collection capacities including strengthening CRVS systems and population-based surveys are key steps to enhancing access to quality data which in turn can contribute to achieving the health-related SDGs.

## Data Availability

The data that support the findings of this study are available on request from the corresponding author.

## Acknowledgements

The views expressed in this paper are those of the authors and do not necessarily reflect those of the World Health Organization. The authors have no financial disclosures or competing interests to report. Ethical approval was not required. All the information is publicly available.

---

**Supplementary Table 1.**
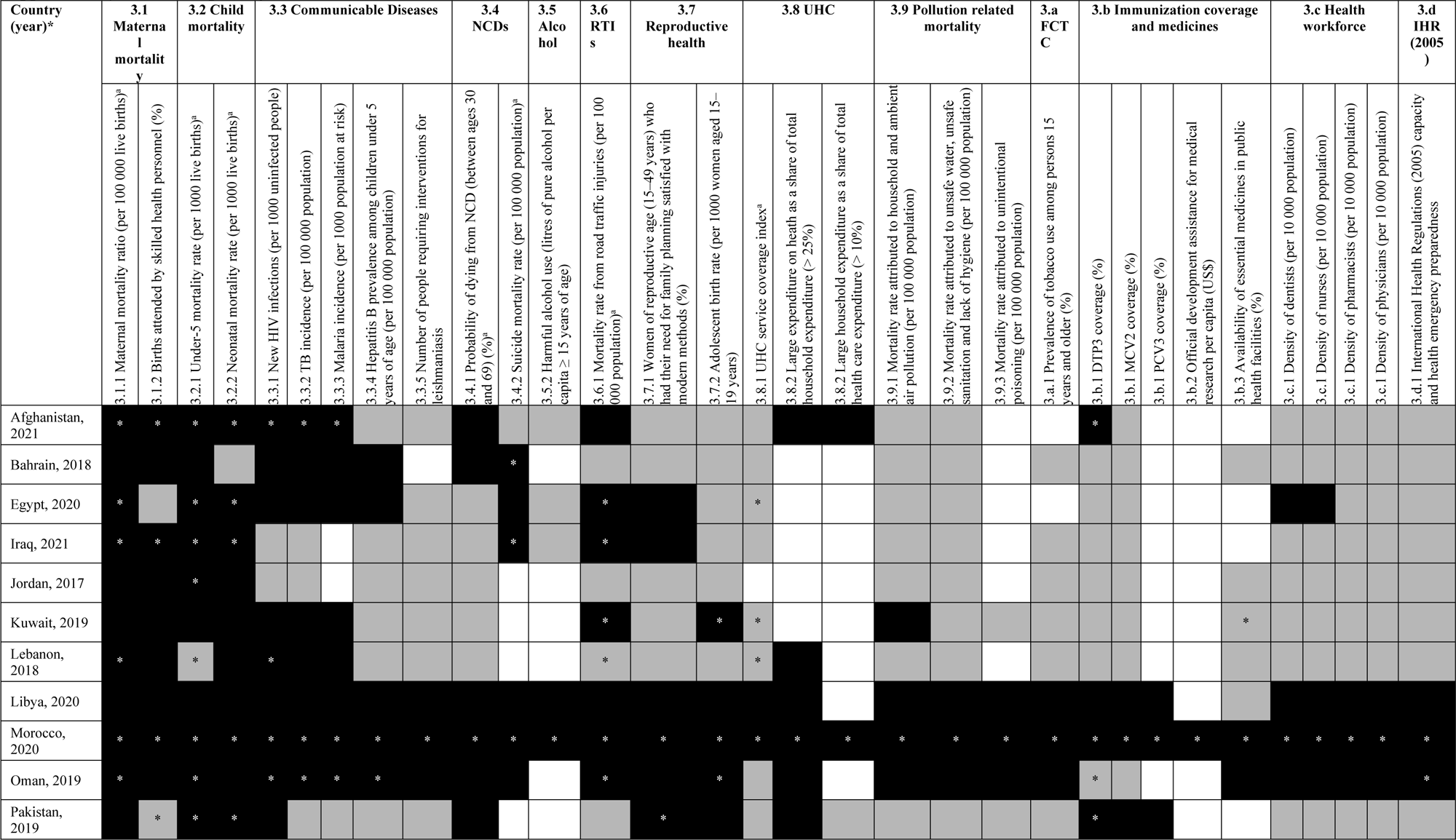
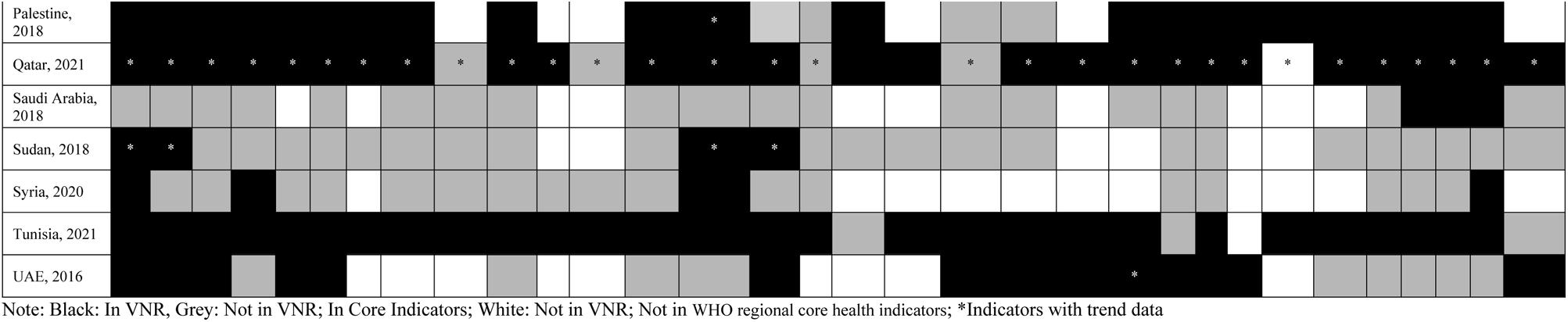
Reporting on SDG3 indicators in the Voluntary National Reviews (VNRs) in the Eastern Mediterranean Region, 2016-2021.

**Supplementary Table 2.**
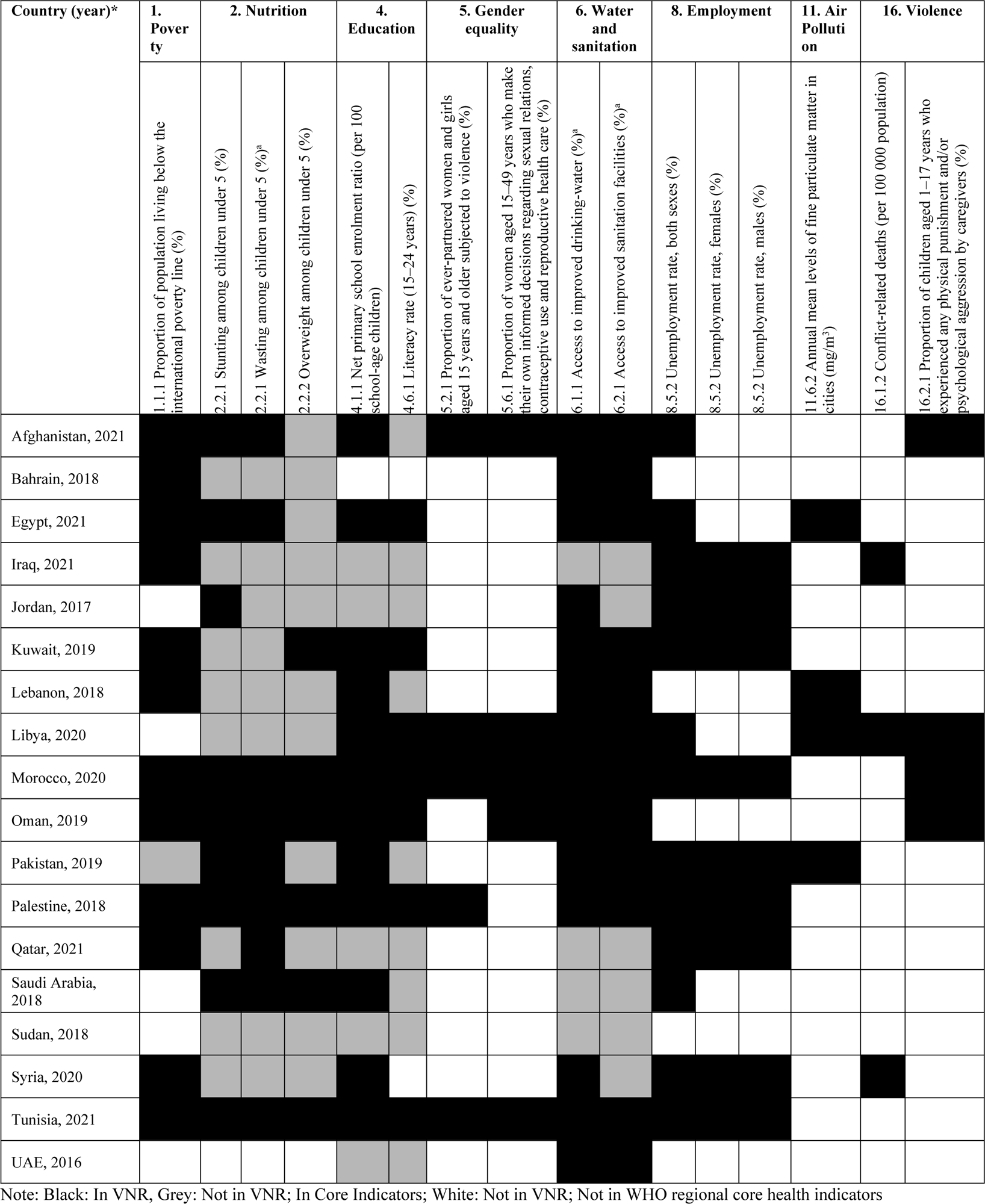
Reporting of health-related SDG indicators in the VNRs.

## Footnotes

a https://sustainabledevelopment.un.org/vnrs/; accessed 10 November 2022.

